# Implementation and evaluation of text message prompts on vaccination uptake: Lessons learned from a large vaccine ecosystem

**DOI:** 10.1101/2025.08.13.25333586

**Authors:** Jacob A. Clarke, Marie Alaghband, Chaz Parkel, Daniel Bridges, Ben Tingey, Emily Price, Ruth Carrico, Wanda Filer

## Abstract

**Introduction:** Vaccines remain one of the most successful public health interventions in the history of medicine. However, recent declines in routine childhood vaccination call for strategies to overcome the many barriers to uptake. This study aimed to implement and assess SMS (Short Message Service) prompting as a tool to improve vaccine uptake within a large vaccine ecosystem.

**Methods:** Utilizing data from the VaxCare ecosystem, a “Vax-to-School” SMS prompt campaign was conducted between May 31 and August 31, 2022, across 1,101 clinics, with 174 clinics participating in the SMS campaign and 927 clinics as control. Eligibility included households with children aged 4-18 years with scheduled clinic appointments. Outcomes included demographic strata, number of appointments and messages, as well as uptake of vaccines under study.

**Results:** A total of 80,205 SMS prompts were sent to 40,018 patients, yielding higher vaccination rates in the SMS+ group (18.8%) compared to the SMS-group (15.9%, P<0.001), though there were fewer appointments in the SMS+ group compared to the SMS-group (mean 1.43 vs 1.51, P<0.001). Significant increases in vaccine administration were observed for Meningococcal B, and Measles Mumps Rubella vaccines, and higher vaccination rates in higher-volume clinics as well as in older children age groups.

**Conclusions:** The study demonstrates that SMS prompts may enhance pediatric vaccine uptake, especially in older pediatric age groups and low-volume program clinics. This intervention holds promise for bolstering immunization coverage across diverse clinic settings and age groups, contributing to public health efforts aimed at reversing the decline in childhood vaccinations.

## Introduction

Routine childhood vaccinations have experienced a significant decline, marking the largest sustained decrease in the last three decades [1,2]. Amidst this decline, it is crucial to identify and address barriers to vaccination. Common parent/ guardian obstacles include challenges in tracking vaccination schedules, lack of understanding about the benefits of vaccination, concerns over potential complications (vaccine hesitancy), and forgetfulness [3,4]. Behavioral prompting through SMS (Short Message Service, commonly referred to as text messaging) has emerged as a strategy to remind individuals regarding the importance of vaccination and, ultimately, improve vaccine uptake [5–7]. SMS prompts involve subtle modifications in information presentation to encourage desired behaviors [7–9]. They can be implemented through interactions between healthcare providers and patients or directly delivered by clinics to patients.

Although the topic of SMS prompting has been explored, there remain several avenues for further investigation, including the exploration of methods to rapidly implement large scale prompting to impact vaccine uptake. The objectives of this study were to: 1) define the processes for implementing an SMS prompt at scale in a large vaccine ecosystem and 2) determine the impact of SMS prompts on childhood vaccine uptake.

## Methods

### The Large Vaccine Ecosystem

This study utilized data from the practice network of a large vaccine ecosystem (VaxCare LLC, Orlando, FL). This ecosystem’s practice network currently includes more than 4,000 active clinical sites of all sizes and patient volumes across most of the United States. The practice network is connected through an end-to-end technology platform that connects vaccine ordering, stocking, inventory management, documentation, tracking, and financial operations. Clinical sites leverage the platform’s capabilities which include obtaining vaccines on consignment, electronic health record system flow integration, and end-to-end life cycle tracking of the vaccine from order to reimbursement. Vaccines are automatically ordered and replenished for clinics, and upon their administration, the platform documents the encounter, bills insurance, and pays the clinic for administering the vaccine to patients. The platform integrates their network to more than 30 electronic health record systems and several state immunization information systems. The platform also allows for crafting and delivery of SMS prompts to patients in conjunction with the scheduled appointments and contact information held in the electronic record system. In the current study, a convenience sample of the total clinical sites were included (n=1,101, 25%).

### Human Subjects Protections

This study was reviewed and approved by the University of Louisville Human Subjects Protection Program Office (IRB #23.0719). All SMS prompts were performed within the guidelines of the Telephone Consumer Protection Act (7 U.S.C. § 227) [10].

### Study Setting

Between 2022 May 31, and 2022 August 31, the vaccine ecosystem launched a “Vax-to-School” vaccination campaign, to include a new SMS prompt strategy. SMS prompts were crafted and dispatched to the parent/guardian of pediatric patients at 174 (approximately 16% of total sites) clinical sites elected to use SMS prompt capability. Although all sites had access to SMS prompts, not all clinics elected to use the capability, allowing for this natural experiment.

### Study Design and Population

To be eligible for the Vax-to-School SMS prompt campaign, patients needed to fulfill the following criteria: (1) households with children aged 4 to 18 years, (2) possession of a valid phone number, (3) a scheduled appointment within the campaign timeframe, and (4) the appointment scheduled at a clinic electing to engage in the campaign.

The study involved two groups of clinics: a group of 174 clinics that elected to engage in the vaccine ecosystem’s Vax-To-School SMS prompt campaign (hereafter noted as SMS+), and a separate group comprising 927 clinics that used the vaccine utilization platform but, did not take part in the SMS prompt campaign (SMS-).

### SMS Prompts

Throughout the campaign, SMS prompts were dispatched to the parent/guardian of children with a valid telephone number and upcoming appointment. The SMS prompt contained the clinic’s contact details, a reminder about the child’s vaccination need, and the option to reply STOP if the recipient wished to opt out of further text communications. Sample SMS prompts are shown in Figure 1. These messages were exclusively sent to the parent/guardian on record for the child and were limited to the child’s first appointment within the campaign period. The SMS prompts were time-triggered, with messages sent 72 and 24 hours before the scheduled clinical appointment, accordingly.

**Figure 1:**
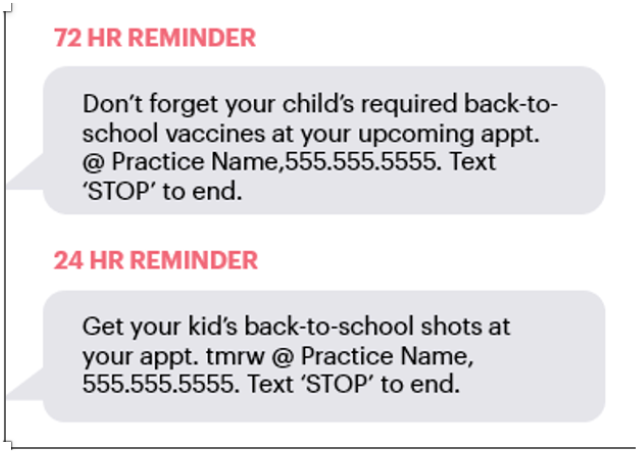
SMS prompt examples.

### Outcome Variables

Patient-level study outcomes included (1) differences in baseline characteristics of patients and clinics, (2) the number of appointments, (3) the number of text messages sent, and (4) vaccine uptake of any non-influenza, non-COVID vaccine during the campaign window.

### Variables

A description of the variables is included in Table 1. Clinics were categorized based on their annual vaccination volume, leading to the distinction of extra-high, high, medium, and low-volume vaccination clinics. The term “extra-high volume” programs pertained to clinics administering over 2,000 annual vaccines, while the “low volume” category encompassed clinics delivering fewer than 400 doses annually.

**Table 1:** Baseline characteristics of pediatric patients (aged 4–18 years old) overall and stratified by SMS prompt status.

|  | Overall | SMS+<sup>1</sup> | SMS-<sup>2</sup> | p-value |
| --- | --- | --- | --- | --- |
| n | 199,734 | 40,018 (20.0) | 159,716 (80.0) |  |
| Sex, n (%) |  |  |  | <0.001 |
| Male | 99,429 (49.8) | 20,357 (50.9) | 79,072 (49.6) |  |
| Female | 100,131 (50.2) | 19,656 (49.1) | 80,475 (50.4) |  |
| Age |  |  |  |  |
| Mean (SD) | 11.46 (4.39) | 10.98 (4.37) | 11.58 (4.39) | <0.001 |
| Age group, n (%) |  |  |  |  |
| 4–6 Years | 41,602 (20.8) | 9,679 (24.2) | 31,923 (20.0) |  |
| 7–10 Years | 45,924 (23.0) | 9,433 (23.6) | 36,491 (22.8) |  |
| 11–12 Years | 30,696 (15.4) | 6,216 (15.5) | 24,480 (15.3) |  |
| ≥13 Years | 81,512 (40.8) | 14,690 (36.7) | 66,822 (41.8) |  |
| Partner vaccination volume, n (%) |  |  |  | <0.001 |
| Extra High | 113,598 (56.9) | 24,268 (60.6) | 89,330 (55.9) |  |
| High | 36,781 (18.4) | 8,247 (20.6) | 28,534 (17.9) |  |
| Medium | 23,507 (11.8) | 5,560 (13.9) | 17,947 (11.2) |  |
| Low | 25,848 (12.9) | 1,943 (4.9) | 23,905 (15.0) |  |
| Created to visit date days |  |  |  |  |
| Median (Q1, Q3) | 17.5 (8.0, 42.0) | 15.5 (7.0, 36.0) | 18.0 (8.0, 42.0) | <0.001 |

<sup>1</sup> SMS+ = clinics participating in SMS prompts<sup>2</sup> SMS- = clinics not participating in SMS prompts

### Statistical Analysis

Baseline characteristics are presented overall and stratified by clinics participating and those not participating in the Vax-To-School program (SMS+ vs SMS-). Frequencies and percentages were used to summarize categorical variables, means with standard deviations (SDs) were used for normally distributed continuous variables, and medians with quartiles (quartile 1, quartile 3; Q1, Q3) were used for non-normally distributed continuous variables. Characteristics were statistically compared between clinic groups using Chi-squared tests for categorical variables, two-sample independent T-tests for normally distributed continuous variables, and Wilcoxon rank-sum tests for non-normally distributed continuous variables. P-values of <0.05 were considered statistically significant. All analyses were performed using Python version 3.9.5 and R version 4.2.3 [11,12].

## Results

### Baseline Characteristics

In this retrospective study, a total of 199,734 patients were included across 1,101 clinics in 26 states. As shown in Table 1, patients had a mean age of 11.46 (SD 4.39); 50.2% (n=100,131) of participants were female and 49.8% (n=99,429) were male. Over half of the patients were part of an extra-high-volume program (56.9%) and the median (Q1, Q3) created to visit date days was 17.5 (8.0, 42.0).

A total of 20% (n=40,018) of patients were in the SMS+ group and 80% (n=159,716) of patients were in the SMS-group. All baseline characteristics were significantly different between SMS+ and SMS-groups.

### Differentials in outcomes by SMS group

The SMS+ group included patients attending clinics across 26 states. A total of 80,205 SMS prompts were sent to a total of 40,018 patients. This equated to an average of 2.00 SMS prompts per patient. The mean number of visits per patient was similar between the SMS+ (1.43 appointments) and SMS-groups (1.51). However, these values were still significantly different (Table 2). Patients in the SMS+ group had significantly higher vaccination rates compared to the patients in the SMS-group (18.8% vs. 15.9%, p<0.001), as seen in Table 2.

**Table 2:** Differentials in clinical outcomes by SMS status.

|  | SMS+<sup>1</sup> | SMS-<sup>2</sup> | p-value |
| --- | --- | --- | --- |
| n | 40,018 | 159,716 |  |
| Text Messages Sent |  |  |  |
| n | 80,205 | – | – |
| Mean per patient | 2.00 | – | – |
| Patients receiving ≥1 vaccination |  |  |  |
| n (%) | 7,512/40,018 (18.8) | 25,322/159,716 (15.9) | <0.001 |
| Appointments |  |  |  |
| n | 57,366 | 241,275 | – |
| Mean (SD) | 1.43 (0.8) | 1.51 (0.9) | <0.001 |
| Patients with > 1 appointment |  |  |  |
| n (%) | 11,839 /40,018 (29.6) | 50,512/159,716 (31.6) | <0.001 |

<sup>1</sup> Clinics participating in SMS prompts<sup>2</sup> Clinics not participating in SMS prompts

When stratified by age group (Table 3) in the SMS-group, vaccine administration was highest in the 11 to 12 years group (32.9%) followed by the ≥13 (16.8%), 4 to 6 (16.1%), and 7 to 10 (2.4%) years age groups. This same rank-order was observed in the SMS+ group.

**Table 3:** Vaccinations by age group.

| Age group | SMS+ | SMS- |
| --- | --- | --- |
| 4-6 Years | 1,622/9,679 (16.8%) | 5,153/31,923 (16.1%) |
| 7-10 Years | 232/9,433 (2.5%) | 869/36,491 (2.4%) |
| 11-12 Years | 2,348/6,216 (37.8) | 8,054/24,480 (32.9%) |
| ≥13 Years | 3,310/14,6890 (22.5%) | 11,246/66,822 (16.8%) |
| Total | 40,018 | 159,716 |

When stratified by program volume (Table 4) in the SMS-group, the vaccination rate was highest among patients associated with extra-high-volume programs (19.9%) followed by those with high (14.6%), medium (11.4%), and low volume programs (5.5%). This differed from patients in the SMS+ group, in which the low-volume programs experienced greater vaccination rates (11.9%) than the medium-volume programs (11.6%).

**Table 4:** Vaccinations by clinic volume.

| Clinic Volume | SMS+ |  | SMS- |  |
| --- | --- | --- | --- | --- |
|  | Number of patients | Vaccinated (%) | Number of patients | Vaccinated (%) |
| Tier 1 (Extra high) | 24,268 | 5,516 (23%) | 89,330 | 17,796 (20%) |
| Tier 2 (High) | 8,247 | 1,119 (14%) | 28,534 | 4,164 (15%) |
| Tier 3 (Medium) | 5,560 | 645 (12%) | 17,947 | 2,037 (11%) |
| Tier 4 (Low) | 1,943 | 232 (12%) | 23,905 | 1,325 (6%) |
| Total | 40,018 | 7,512 | 159,716 | 25,321 |

### Effects on Vaccine Uptake

When stratifying by the vaccine administered, administration of all vaccines was increased in the SMS+ group compared to the SMS-group, though not all differences were statistically significant. Table 5 shows vaccinations by SMS group with significance estimates. Specifically, greatest percent increase in vaccine administration occurred with Meningococcal B (MenB) (64.2%), followed by Measles Mumps Rubella (MMR) (36.2%). The greatest administration increases per 10,000 patients occurred with MenB (Proportion difference per 10,000 [95% CI]: 162.1 [141.1–183.0]), followed by HPV9 (95.1 [68.3-–121.8]). Table 5 shows vaccines by SMS group.

**Table 5:** Vaccines by SMS group.

|  | Vaccinations per 10,000 patients |  | Absolute rate difference (95% CI<sup>3</sup>) | Relative difference | General age groups |
| --- | --- | --- | --- | --- | --- |
|  | SMS+<sup>1</sup> | SMS-<sup>2</sup> |  |  |  |
| Men ACWY | 806.89 | 715.58 | 91.3 (61.8, 120.8) | 12.8 | 11–12; ≥13 |
| IPV | 35.98 | 31.87 | 4.1 (–2.4, 10.6) | 12.9 | 4–6 |
| Hepatitis A | 80.71 | 71.31 | 9.4 (–0.3, 19.1) | 13.2 | 0–4 |
| Tdap | 476.29 | 409.04 | 67.3 (44.2, 90.3) | 16.4 | 7–10 |
| DTaP | 28.24 | 24.23 | 4.0 (–1.7, 9.7) | 16.5 | 4–6 |
| HPV9 | 655.7 | 560.62 | 95.1 (68.3, 121.8) | 17.0 | 7–10; 11–12; ≥13 |
| IPV+DTaP | 308.61 | 251.38 | 57.2 (38.6, 75.8) | 22.8 | 4–6 |
| MMRV | 241.64 | 192.22 | 49.4 (32.9, 65.9) | 25.7 | 4–6 |
| Varicella | 125.44 | 94.42 | 31.0 (19.1, 42.9) | 32.9 | 4–6 |
| MMR | 120.2 | 88.28 | 31.9 (20.3, 43.5) | 36.2 | 4–6 |
| Men B | 414.31 | 252.26 | 162.1 (141.1, 183.0) | 64.2 | 7–10; 11–12; ≥13 |

<sup>1</sup> SMS+ clinics participating in SMS prompts
<sup>2</sup> SMS- clinics not participating in SMS prompts
<sup>3</sup> Confidence interval

## Discussion

Our study supports the use of SMS prompts as a strategy for increasing pediatric vaccine uptake. In our secondary analysis, the utilization of SMS prompts was associated with an increase in pediatric vaccination rates. Notably, the impact of these SMS prompts was more pronounced among older pediatric patients. SMS prompts were also associated with increased administration rates of MenB and Human Papillomavirus-9 (HPV9) vaccines. Additionally, the impact of SMS prompts was highest for patients in low-volume programs. Our findings also harmonize with recent research exploring the influence of SMS prompts on vaccination rates [5,7,8,13–20]. Further, the SMS prompt strategy aligned with successful strategies from previous research, such as employing multiple SMS prompts, utilizing professional language that fosters ownership, and timing messages closer to clinical visits [5,13,14,21].

Our study had other interesting finds involving clinic vaccination program size. Clinics linked to low-vaccination-volume programs exhibited the most substantial increase in vaccination rates between the SMS+ and SMS-groups. This trend might be attributed to the resource constraints that often challenge these smaller practices. The strains on primary care during the COVID-19 pandemic, including staffing limitations and reduced in-person visits, further compounded vaccination challenges. SMS prompts were designed to support clinical vaccination programs without burdening the staff, potentially bridging the gap in vaccination reminders due to limited staffing. Further, the ability to craft and deploy SMS prompts independent of an electronic medical record provides opportunities for organizations using systems that lack messaging functionality. Future studies could delve into the effects of such vaccine management and communication strategies on program scalability, as well as population or vaccine focus.

Another interesting finding of this study spotlighted MenB as the vaccine with the largest absolute and relative increase in administration rate. This observation could be attributed to MenB’s status as a vaccine subject to shared decision-making. The SMS prompts might have prompted enhanced shared clinical decision-making conversations with healthcare providers, leading to increased uptake. Notably, the Measles, Mumps, Rubella, and Varicella (MMRV) vaccine and HPV9 vaccine displayed the second-highest relative and absolute increases in administration rates, respectively. Both vaccines are of heightened public concern due to declining administration rates and the imperative to address care gaps and minimize patient vulnerability to preventable diseases [22]. Our findings underscore the use of SMS prompts as a potential remedy for reversing the decline in these at-risk vaccine rates. Subsequent studies should explore the public health implications of SMS prompts’ role in boosting uptake rates of vaccines facing declining rates.

Vaccine administration rates are influenced by complex factors including a clinic’s in-house availability of specific vaccines, as well as operational resources. In our study, offices associated with small vaccination volume programs saw the greatest increase in vaccination rates between intervention and control. This may be because these lowest volume vaccinators logically have the fewest staff and other resources. Primary care practices were particularly challenged during the pandemic, with staffing constraints and fewer in person visits. Program nudges were designed to support vaccination programs in the office without burdening the practice staff with extra work. From a behavioral perspective, ‘nudges’ are methods that can be used to present, frame, or craft messages or information in order to influence behavior while maintaining freedom of choice [9]. Consequently, text message nudges may have helped fill the gap in vaccination reminders that could result from low staffing by systematically offloading of this work to the vaccine provider. Further, use of media and behavioral sciences interventions could be an important element in the multipronged strategy supporting efforts to improve vaccine acceptance and vaccine schedule adherence, even if incremental improvements are small. In addition, traditional barriers like up-front vaccine administration costs, such as stocking and reordering, can hinder clinics from offering certain vaccines. In our study, the vaccine ecosystem capabilities may have mitigated these barriers, ensuring uniform access to vaccines across clinics.

Another intriguing finding is the significant association between SMS prompts and vaccine uptake among older pediatric age groups, particularly those aged 13 and above. Adolescent vaccines are well known for needing more attention since adolescents do not have the same frequency of recommended office visits as younger children and often do not keep well checks, instead being seen for camp and state-required school physicals [23]. Our results point to a pivotal opportunity for closing care gaps in this harder-to-reach population.

Our findings are of particular importance as little has been written regarding the impact of text messaging as an influencer to improve vaccination rates among school-aged children and the vaccines necessary for school attendance. Since the SARS-CoV-2 pandemic, relevant information in the literature with few exceptions has involved impact in countries other than the US [5,24–27]. One recent study focused on US health systems studied only COVID-19 vaccination [20]. To that end, identifying the impact text messaging may have among US parents and children makes continued exploration of this approach an important public health consideration.

### Strengths and Limitations

In addition to new information regarding the impact of text messaging on school-aged vaccinations in the US, our study’s expansive scale is a strength, surpassing prior investigations regarding the volume of messages sent, the number of patients involved, and geographical representation across the US. This scale makes the results more likely to generalize the wider US pediatric population.

Alongside the aforementioned strength, this study has several limitations. First, all patients in the vaccine ecosystem are commercially insured, making it difficult to compare differences among those in the US Vaccines for Children Program or when seeking to improve vaccination among uninsured or underinsured populations. Second, SMS prompts were sent to phone numbers without verification of their capability to receive text messages. Consequently, there exists the possibility that certain patients did not receive these messages, introducing an element of potential non-receipt bias. Third, our dataset lacked information on patient “no-show” instances or vaccinations obtained outside the scope of this study, potentially leading to a misclassification bias. We hypothesize that incorporating these data could have yielded even more pronounced vaccine rate increases within the SMS+ group. Fourth, we did not differentiate between patients scheduled solely for a vaccine visit and those who were vaccinated as part of an existing or new appointment and included all of those visits as a vaccinating opportunity. This choice may have impacted the denominator and therefore impacted results. Further, since this study was not randomized and the statistical analysis was unadjusted, confounding bias may play a role in the interpretation of results.

Our study encompassed a diverse range of clinics, yet it did not explore regional or clinic-specific variations beyond program vaccination volume. Furthermore, the inclusion of clinics into the SMS+ group was contingent on their voluntary opt-in for receiving text messages, possibly engendering selection bias.

## Conclusions

In this study, the implementation of SMS prompts, tailored for their target audience, content, and timing, demonstrated an association with heightened vaccination rates for non-flu, non-COVID vaccines in pediatric patients. Our study underscores the utility of SMS prompts in influencing pediatric vaccine uptake. These findings hold potential for public health efforts to enhance immunization coverage in diverse clinic settings, age groups, and vaccine types.

## Declarations

## Ethics approval and consent to participate

This study was reviewed and approved by the University of Louisville Human Subjects Protection Program Office (Protocol #23.0719). All SMS prompts were performed within the guidelines of the Telephone Consumer Protection Act [10]. All experiments were performed in accordance with relevant guidelines and regulations (such as the Declaration of Helsinki). The requirement for informed consent was waived by the University of Louisville Human Subjects Protection Program Office because of the retrospective nature of the study.

## Consent for publication

Not applicable.

## Data Availability Statement

De-identified data presented in this study are available on reasonable request from the corresponding author. The data were collected as part of VaxCare business practices and are not publicly available due to patient privacy.

## Author Contributions

JC, MA, CP, DB, BT, EP were responsible for primary manuscript development. RC was responsible for data review and manuscript editing. WF was responsible for critical review of the manuscript and final editing. All authors were responsible for final manuscript review and approval.

## Acknowledgments

The authors express appreciation to the clinic personnel who work to improve vaccination rates among their patients and for their participation in this text messaging campaign.

